# Assessing Causal Relationships Between Diabetes Mellitus and Idiopathic Pulmonary Fibrosis

**DOI:** 10.1101/2024.01.04.24300827

**Authors:** Samuel Moss, Cosetta Minelli, Olivia Leavy, Richard Allen, Nick Oliver, Louise Wain, Gisli Jenkins, Iain Stewart

**Author notes:** Corresponding author: Mr. Samuel Moss, Margaret Turner Warwick Centre for Fibrosing Lung Disease, National Heart and Lung Institute, Imperial College London, London, UK**. Orcid: 0000-0002-1872-9110.**. **Author Contributions:** All authors contributed to analysis, drafting, and final approval of this manuscript.

## Abstract

**Rationale:** Idiopathic Pulmonary Fibrosis (IPF) is a disease of progressive lung scarring. There is a known association between IPF and diabetes mellitus (DM), but it is unclear whether this association is due to causal relationships between these traits.

**Objectives:** To examine causal relationships between DM, diabetes-associated traits, and IPF using a Mendelian randomisation approach.

**Methods:** Following a two-sample Mendelian randomisation (MR) approach, we used genetic variants identified from genome wide association studies (GWAS) for type 1 diabetes (T1D), type 2 diabetes (T2D), glycated haemoglobin level (HbA1c), fasting insulin level, and body mass index (BMI) to assess for evidence of causal effects of these traits on IPF risk. Further analyses using pleiotropy-robust and multivariable MR methods were performed to account for the inherent complexity of the traits being investigated.

**Results:** Results did not suggest that either T1D (OR = 1.00, 95% CI: 0.93-1.07, p = 0.902) or T2D (OR = 1.02, 95% CI: 0.93-1.11, p = 0.692) are in the causal pathway of IPF. No significant effects were suggested of HbA1c (OR = 1.19, 95% CI: 0.63-2.22, p = 0.592) or fasting insulin level (OR = 0.60, 95% CI: 0.31-1.15, p = 0.124) on IPF risk, but effects of BMI on IPF risk were indicated (OR = 1.44, 95% CI: 1.12-1.85, p = 0.004).

**Conclusion:** This study suggests that DM and IPF are unlikely to be causally linked. This comorbid relationship may instead be driven by shared risk factors or treatment effects.

**Key messages:** **What is already known on this topic:** Idiopathic pulmonary fibrosis is associated with diabetes mellitus epidemiologically, but it is unclear if these traits are linked by causal effects.

**What this study adds:** Idiopathic pulmonary fibrosis and diabetes mellitus are unlikely to be causally linked, suggesting that shared environmental risk factors or treatment effects may drive this comorbid relationship.

**How this study might affect research, practice, or policy:** Further research investigating the relationship between diabetes mellitus and idiopathic pulmonary fibrosis should focus on potential shared risk factors such as smoking, and treatment effects including corticosteroid use.

## Introduction

Idiopathic pulmonary fibrosis (IPF) is a chronic disease characterised by progressive and uncontrolled scarring of the lung. The development of fibrosis in IPF reduces lung function over time and is typically fatal within 3-5 years of diagnosis^1,2^. The mechanisms underlying IPF risk and progression are not fully understood.

Diabetes mellitus (DM) is a group of diseases characterised by hyperglycemia due to disrupted insulin production, reduced effectiveness of secreted insulin, or a combination of both^3^. Over 400 million people have DM globally and incidence has been shown to be increasing^4^. Type 1 diabetes is caused by autoimmune destruction of pancreatic beta cells and subsequent insulin deficiency^5^. Type 2 diabetes (T2D) is a long-term condition characterised by insulin resistance, dysregulation of blood glucose levels, and a progressive decline in beta cell function^6^. Studies show increased prevalence of DM in IPF patients when compared with controls^7–9^ and comorbid DM has been shown to be associated with increased mortality in IPF patients^10^. As current studies assessing this association have primarily used mixed DM cohorts it is unclear if prevalence of DM in IPF patients varies with DM subtype. Furthermore, decreased lung function has been reported in DM cohorts and is correlated with having glucose above target^11–13^.

The potential mechanisms by which DM may be associated with IPF risk are complex. Hyperglycemia has been linked to immune dysfunction, chronic low-grade inflammation, increased infection rate^14,15^, and worse immune-related clinical outcomes in critically ill patients^14–16^, with outcomes improving with optimised glycemic control^16–18^. Immune dysregulation has a key role in fibrogenesis^19,20^; similarly, hyperglycemia may contribute to IPF risk through impaired DNA repair mechanisms and subsequent increase of cellular senescence^21–23^. The potential relationship between T2D and IPF is further complicated by metabolic factors becoming dysregulated both due to T2D and during disease development. Factors such as chronic hyperglycaemia, obesity, and hyperinsulinemia contribute to the development of T2D^24–26^ and may therefore independently drive IPF pathogenesis (Figure 1).

**Figure 1:**
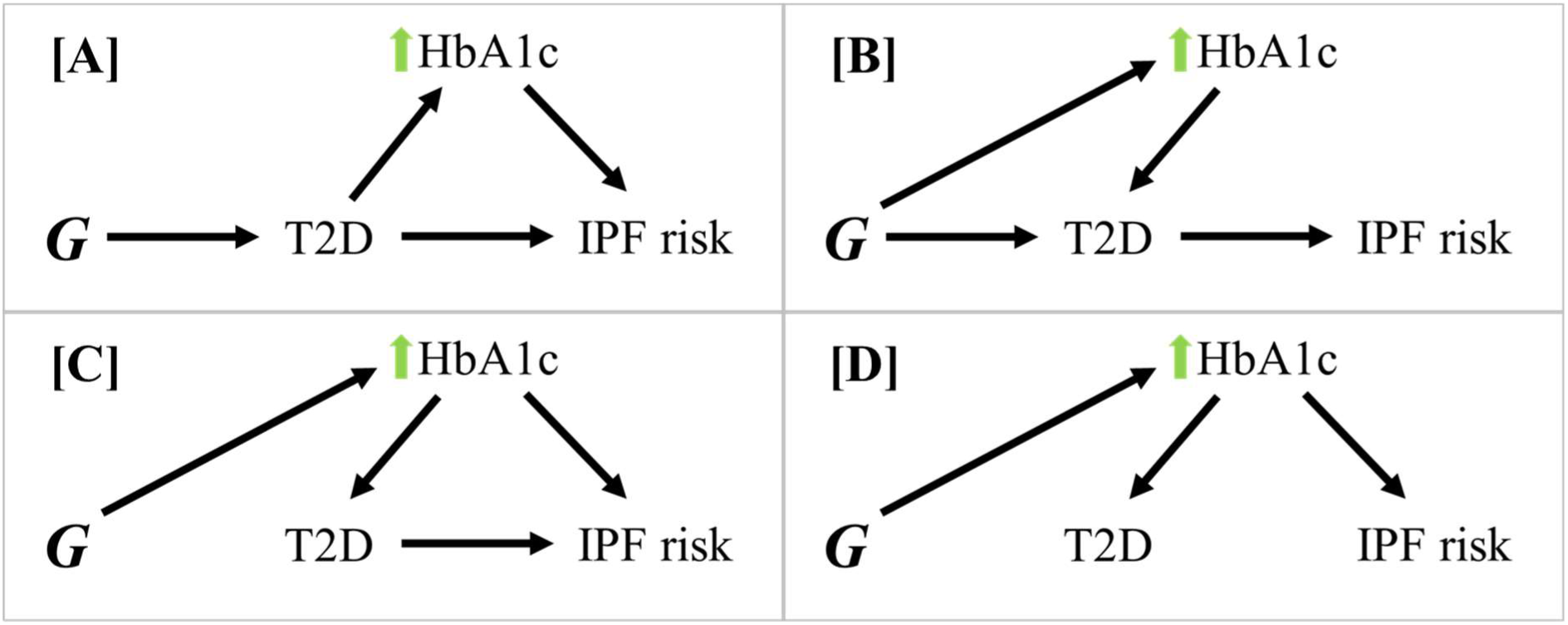
Directed acyclic graphs (DAG) showing potentially interacting pathways of effect on IPF risk. Let *G* be the genetic instrument being used to estimate causal effect on IPF risk. [A] Any effects of *G* on the causal pathway between T2D and IPF (such as raised HbA1c downstream of T2D) act through T2D, therefore not resulting in confounding effects. [B] Any pleiotropic effects on HbA1c do not influence IPF risk. Effects through this pathway are valid, but the continuous HbA1c exposure may be a stronger instrument for this effect [C]. Effects of *G* on T2D acting through HbA1c also increase IPF risk independently of T2D (horizontal pleiotropy) and invalidate the independence assumption. [D] Effects of *G* on T2D act through HbA1c, but HbA1c influences IPF risk independently of T2D, also invalidating the independence assumption.

Mendelian randomisation (MR) studies aim to assess the extent of causal relationships between modifiable disease risk factors or exposures and a disease or disease-related outcome. MR methods typically use germline genetic variants as instruments for the exposure of interest, therefore allowing for control of environmental effects that could confound observational studies. Effective causal estimation and control of confounding requires genetic instruments that are: (1) reliably associated with the exposure; (2) only associated with the outcome through the exposure; (3) independent of confounders that influence both the exposure and outcome^27–30^. Horizontal pleiotropy, the phenomenon of genetic instruments influencing the outcome through pathways independent of the exposure of interest^31^, can confound causal estimation in MR and violates assumptions (2) and (3).

The aim of this study was to estimate the causal effect of both T1D and T2D (exposure) on IPF risk (outcome) using a two-sample MR approach. Additional univariable and multivariable MR analyses were performed to assess the causal effects on IPF risk of continuous exposures representing main factors associated with DM – Raised blood glucose (raised HbA1c level), dysregulated insulin (raised fasting insulin level), and obesity (raised BMI). As a secondary analysis, we also investigated the presence of a causal effect in the opposite direction, that is the effect of IPF on DM risk (Supplementary Analyses 1 and 2).

## Methods

### Study populations

Genetic association statistics were retrieved from genome-wide association studies (GWAS) of T1D^32^ (6,683 European ancestry cases, 12,173 European ancestry controls, 2,601 European ancestry affected sibling pair families, 69 European ancestry trios), T2D^33^ (61,714 European ancestry cases, 1,178 Pakistani ancestry cases, 593,952 European ancestry controls, 2,472 Pakistani ancestry controls), IPF risk^34^ (4,125 European ancestry cases, 20,464 European ancestry controls), HbA1c level^35^ (sample size: 88,355 of European ancestry), fasting insulin level^36^ (sample size: 151,013 of European ancestry), and BMI^37^ (sample size: 681,275 of European ancestry). A secondary analysis of T2D on IPF was performed using data from a GWAS of only European ancestry participants^38^ (12,931 European ancestry cases, 57,196 European ancestry controls) (Supplementary Analysis 1). An additional sensitivity analysis of HbA1c on IPF used a subset of functionally glycemic (associated with glycemic traits as defined by Wheeler *et al*. (2017) ^35^) HbA1c variants (supplementary analysis 2).

No participants in the HbA1c or fasting insulin GWAS cohorts had a diagnosis of diabetes.

### Genetic instrument selection

Genetic instruments were required to be associated with each exposure at genome-wide significance level (p<5×10^-8^). A clumping approach was used to select independent signals (r^2^ < 0.001 within a 10,000 kb window) using the 1000 Genomes Project reference panel through *ieugwasr*^39^. All genetic variants were required to be sufficiently strong instruments (F>10) for each exposure to avoid weak instrument bias. Any missing variants in the outcome datasets were replaced with suitable proxies (R^2^>0.7) where available, using *LDlinkR*^40^.

### Statistical analyses

A random-effects inverse-weighted (IVW-RE) MR method^41,42^ was used to estimate the overall effect of each exposure on outcomes using the *MendelianRandomization* R package. MR-Egger^43^ and weighted median^44^ (MR-WM) MR methods were used here to provide pleiotropy-robust causal estimates in a sensitivity analysis. MR-Egger uses meta-regression to estimate causal effects adjusted for directional pleiotropy, whereas MR-WM allows for pleiotropic effects in up to 50% of instruments. MR-PRESSO^45^ and MR-Lasso^46^ extensions of the IVW method were used to identify and remove likely pleiotropic genetic instruments. MR-PRESSO identifies likely pleiotropic instruments through variant contribution to heterogeneity in the IVW meta-analysis of variant-specific MR estimates, whereas MR-Lasso includes an intercept term to estimate genetic effects bypassing the risk factor of interest. All univariable MR methods, other than MR-PRESSO, have multivariable equivalents that were applied in MVMR (MVMR-IVW^47,48^, MVMR-Median^49^, MVMR-Egger^50^, MVMR-Lasso^49^).

## Results

### Selection of genetic instruments

Thirty-five genetic variants were selected as instruments for T1D, and 118 genetic variants were selected as instruments for T2D. Genetic instrument sets were similarly identified for HbA1c level (37 variants), fasting insulin (36 variants), and BMI (131 variants). All selected variants for these exposures had corresponding GWAS summary results for IPF.

### Estimation of causal effects

No significant causal effect was suggested of T1D on IPF risk (IVW-RE, Odds Ratio (*OR*) = 1.00, 95% Confidence Interval (*95% CI*): 0.93-1.07, p = 0.902). MR estimates similarly did not suggest significant causal effects of T2D on IPF risk (IVW-RE, *OR* = 1.02, *95% CI*: 0.93-1.11, p = 0.692). While there was statistically significant heterogeneity present analyses of T1D on IPF (Q = 65.56, p < 0.01, I^2^ = 48.1%), and of T2D on IPF (Q = 148.43, p = 0.03, I^2^ = 21.2%), results from pleiotropy-robust MR methods were consistent with those of IVW-RE (Figure 2 and Figure 3). Further, removal of likely pleiotropic variants reduced heterogeneity scores in analyses of T1D on IPF (MR-Lasso, Q = 33.22, p = 0.360, I^2^ = 6.7%), and T2D on IPF (MR-Lasso, Q = 105.43, p = 0.605, I^2^ = 0%), but MR estimates were consistent with the IVW-RE approach.

**Figure 2:**
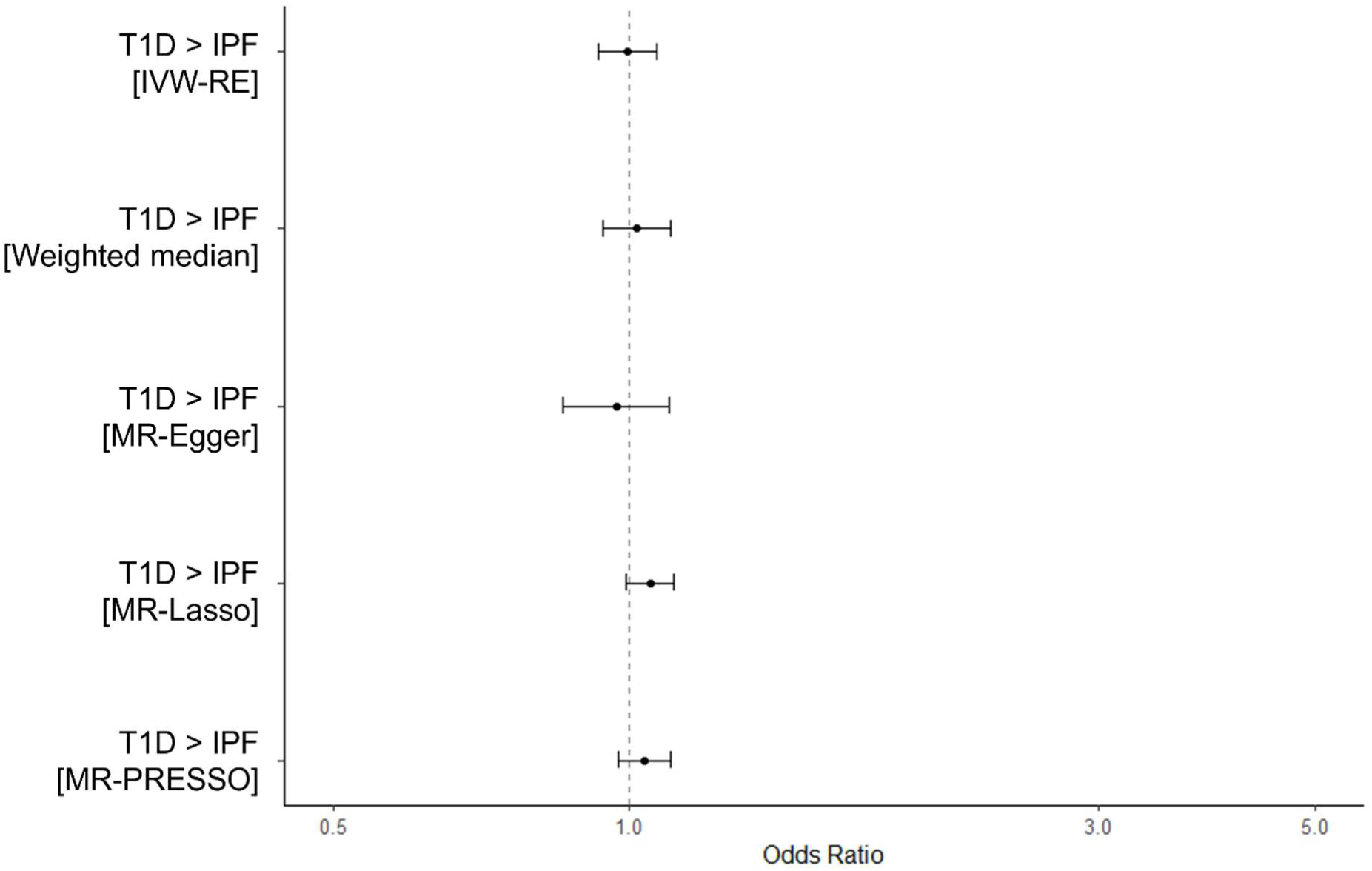
Overall causal estimates of T1D on IPF risk estimated using different MR approaches. Error bars show 95% confidence intervals.

**Figure 3:**
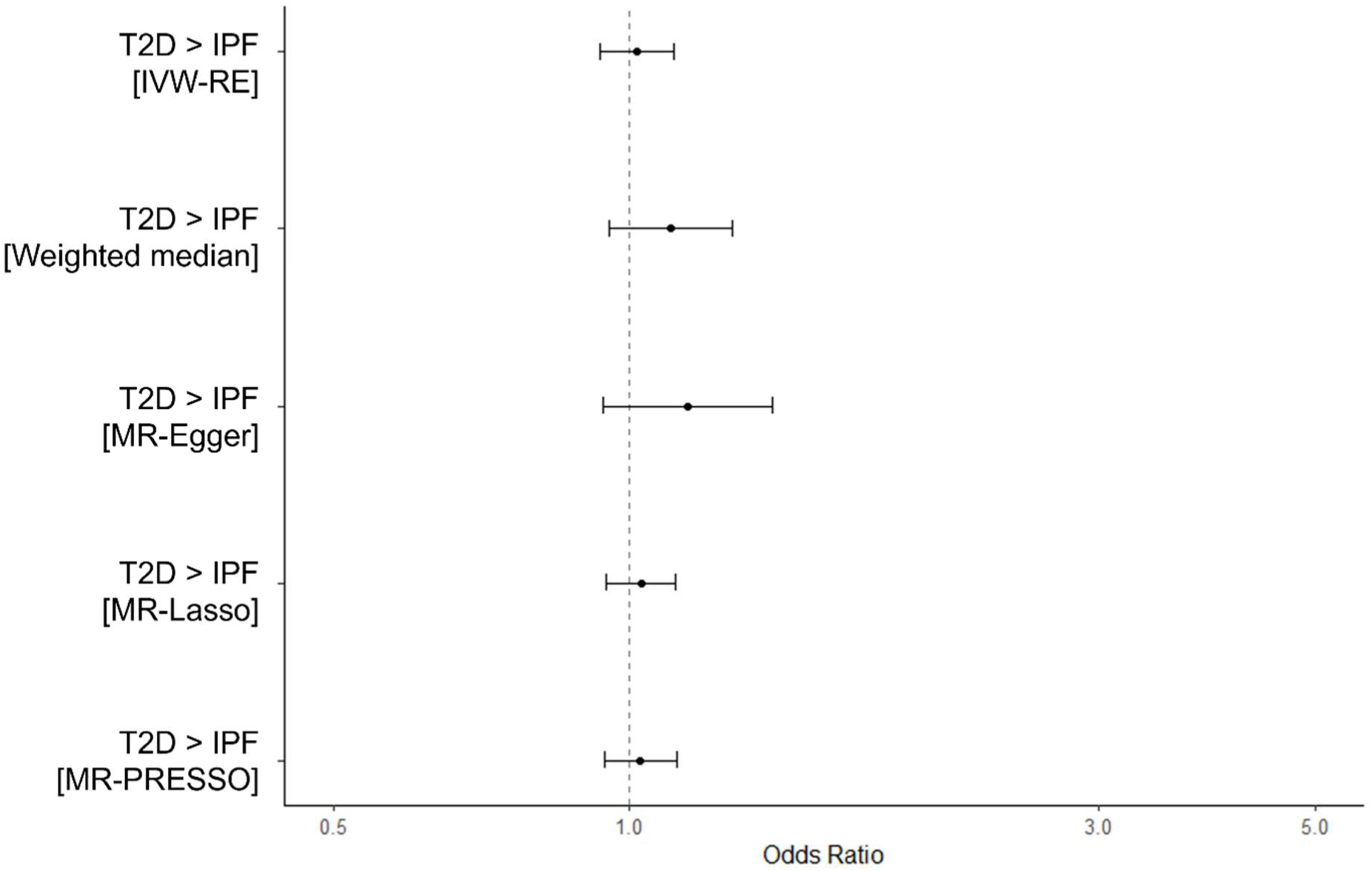
Overall causal estimates of T2D on IPF risk estimated using different MR approaches. Error bars show 95% confidence intervals.

No causal effect of IPF on T2D (Supplementary Analysis 3) or T1D (Supplementary Analysis 4) was identified when the MR approach was applied in this direction.

### Estimation of causal effects using continuous exposures

We found no evidence suggesting that genetically proxied raised HbA1c causes IPF (IVW-RE, OR = 1.19, 95% CI: 0.63-2.22, p = 0.592). Results of MVMR analyses did not suggest any independent effects of T2D (MV-IVW, OR = 1.03, 95% CI: 0.92-1.15, p = 0.601) or raised HbA1c (MV-IVW, OR = 1.14, 95% CI: 0.52-2.52, p = 0.746) on IPF risk. Estimates calculated using pleiotropy-robust methods were consistent across univariate and MVMR analyses (Figure 4A and Figure 5A). Outcomes from the MR analysis of HbA1c on IPF using a subset of only glycemic HbA1c variants were consistent with outcomes of main HbA1c analyses (Supplementary Analysis 2).

**Figure 4:**
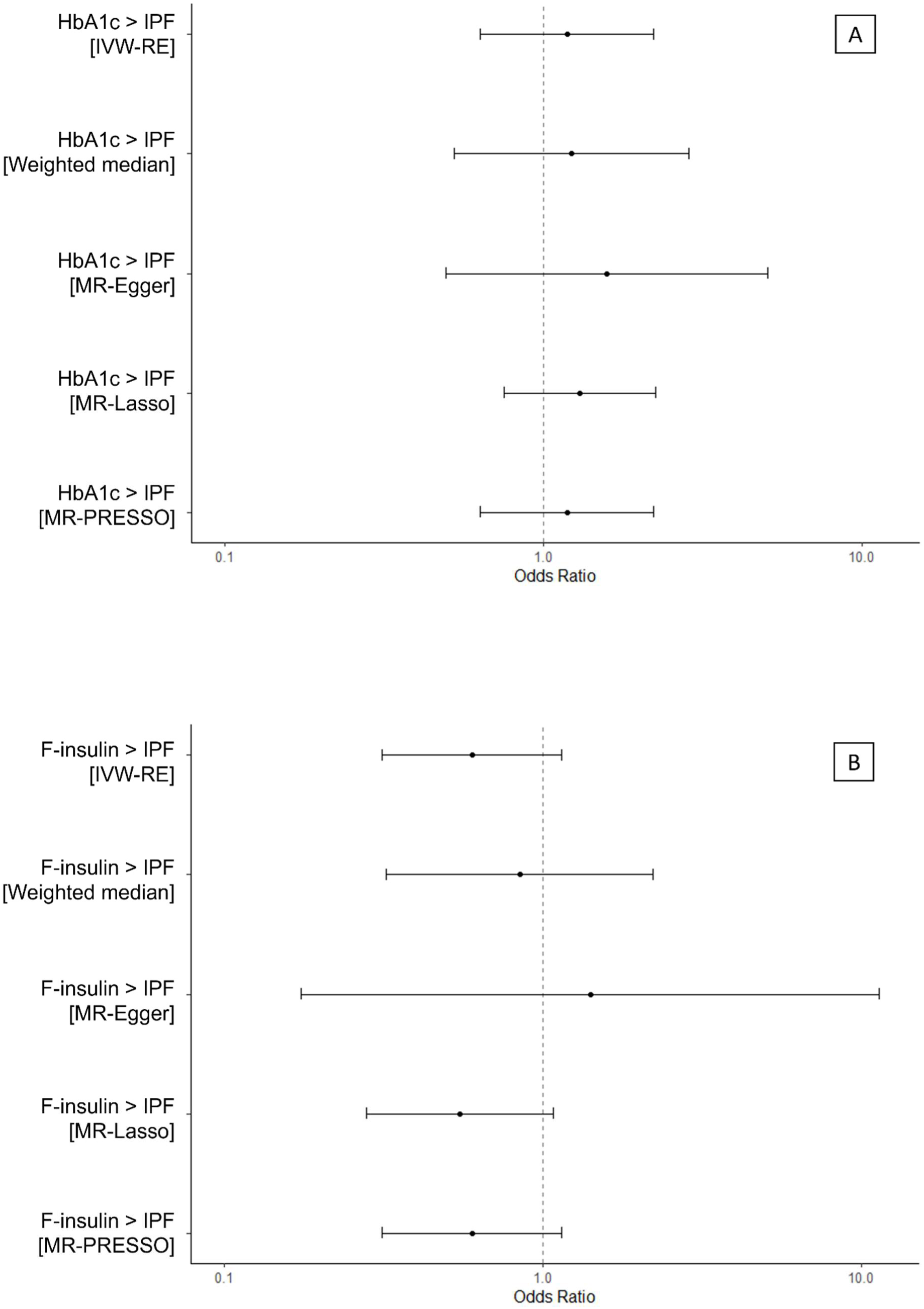
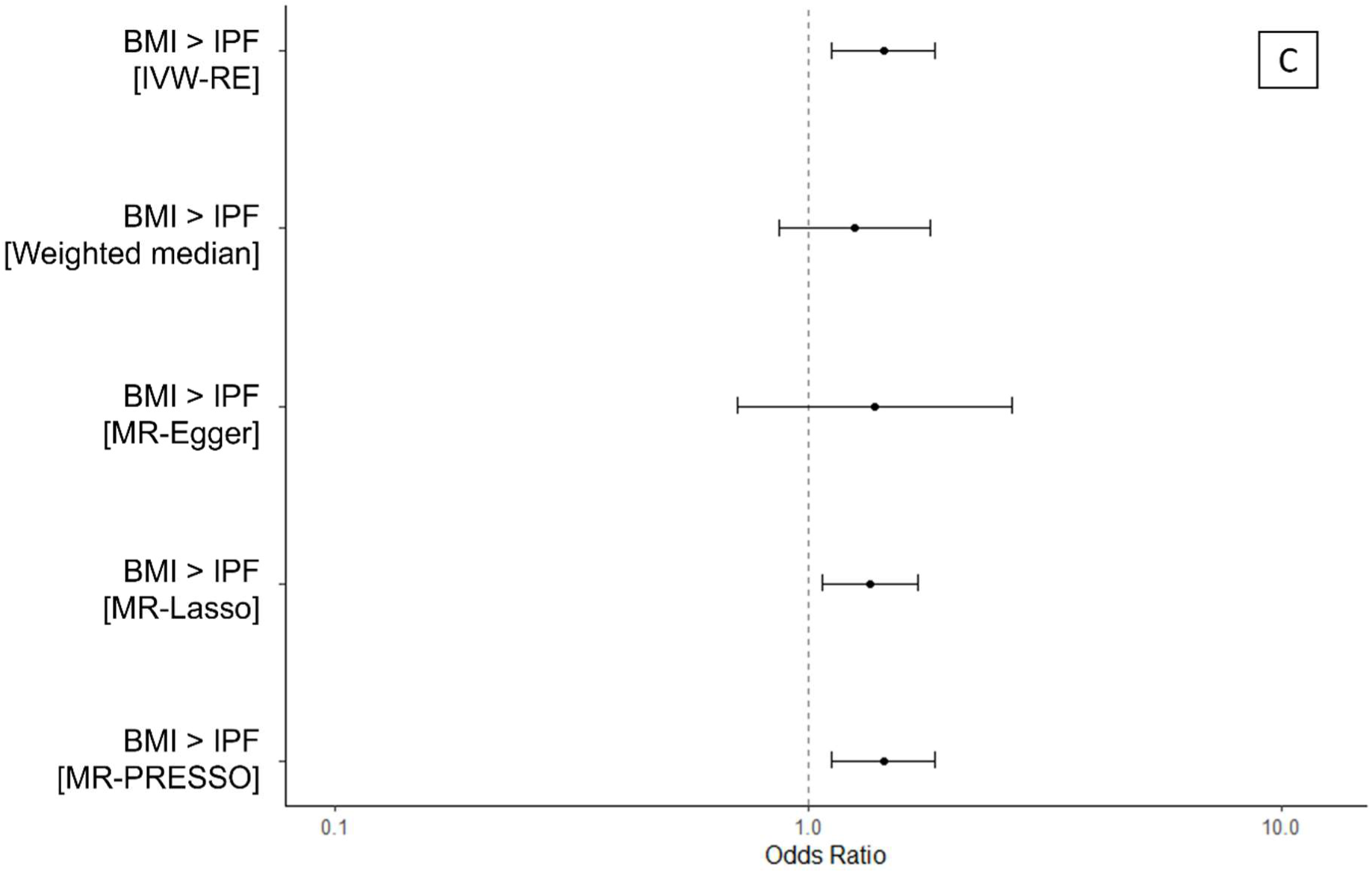
Causal estimates from univariable MR analyses of T2D-associated continuous exposures on IPF risk. [**A**] Causal estimates of genetically proxied raised HbA1c level on IPF risk. **[B]** Causal estimates of genetically proxied raised fasting insulin (F-insulin) level on IPF risk. **[C]** Causal estimates of genetically proxied raised BMI on IPF risk. Odds ratios were calculated from beta estimates. Error bars show 95% confidence intervals.

**Figure 5:**
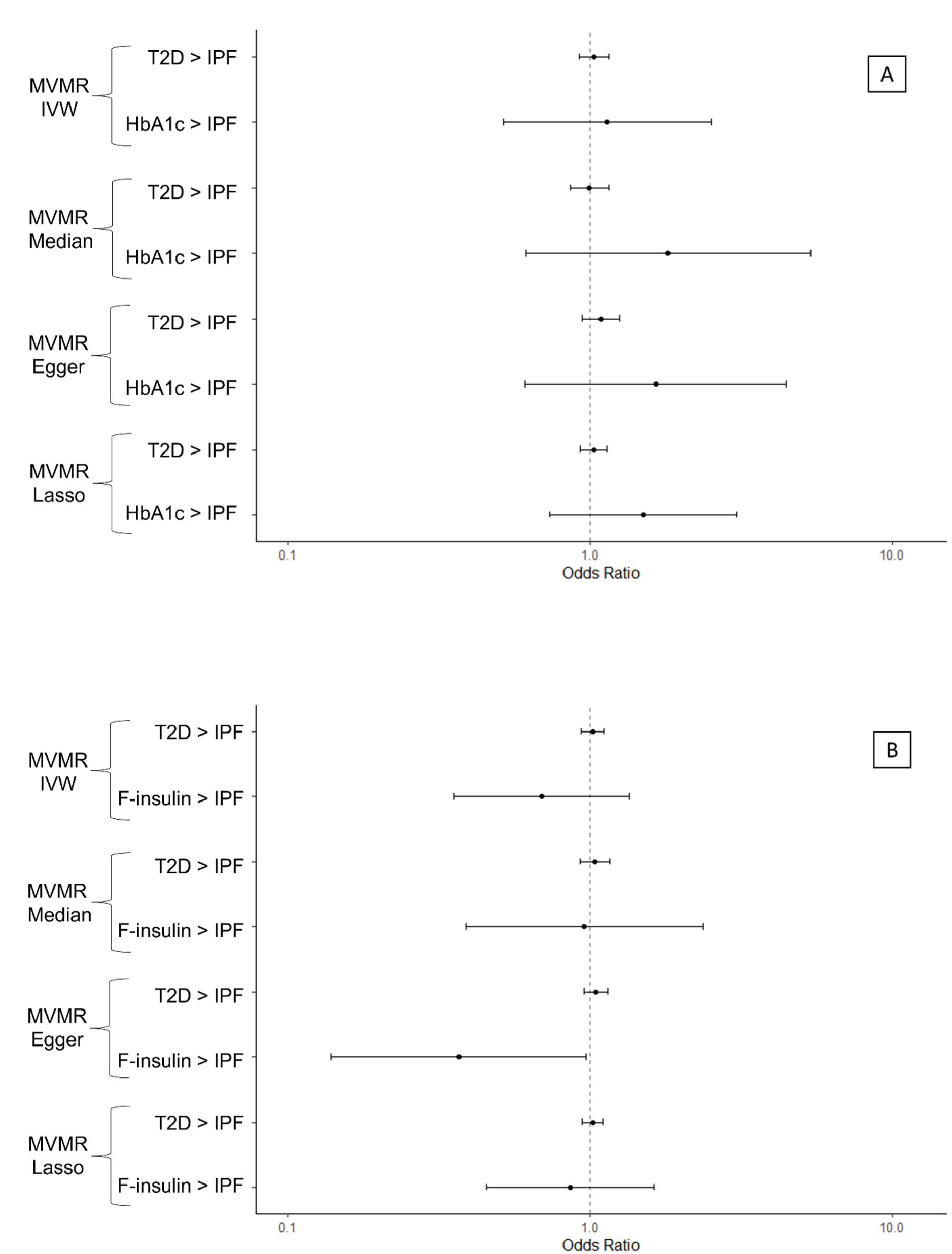
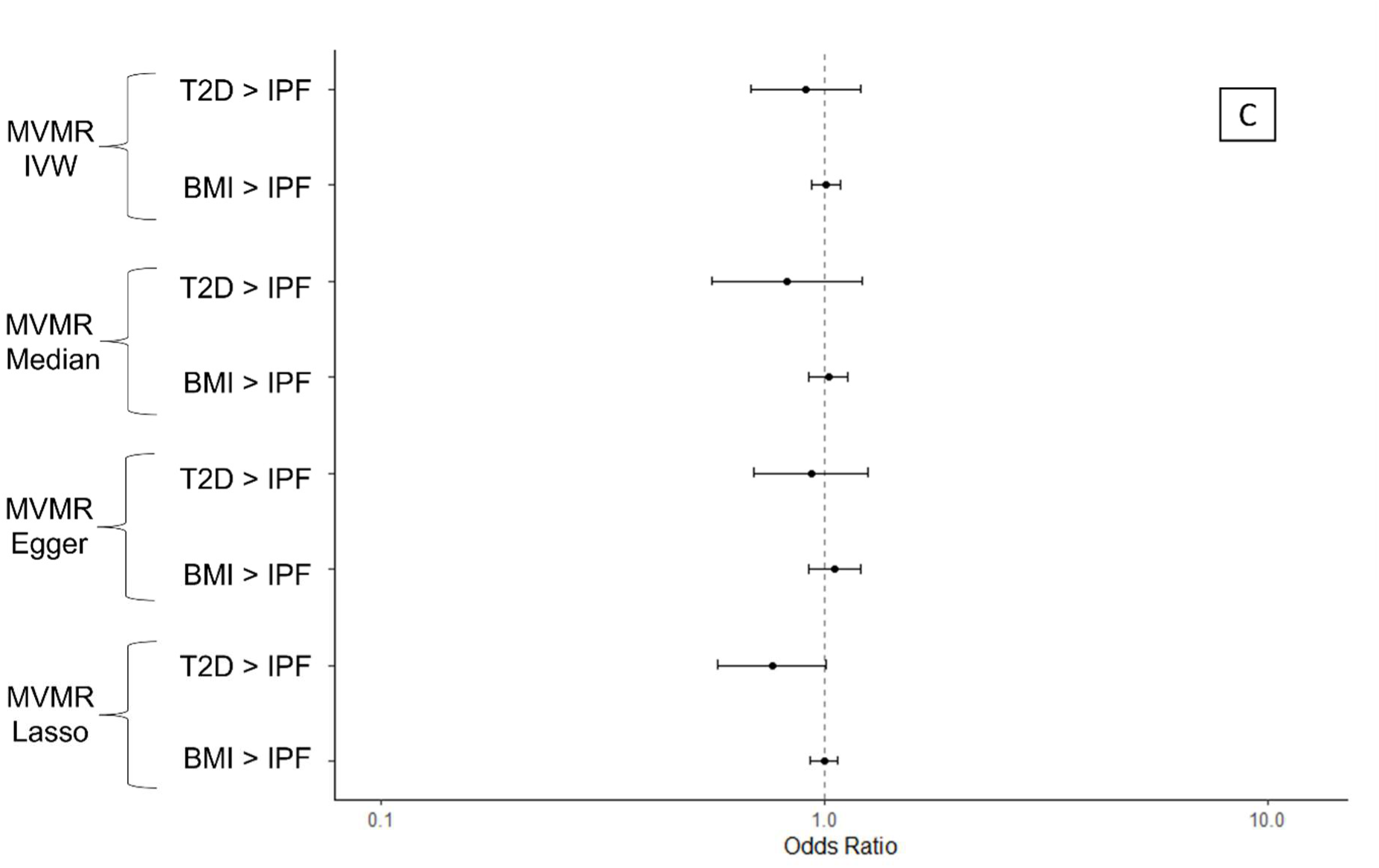
Causal estimates from multivariable MR analyses of T2D, and T2D-associated continuous exposures on IPF risk. [**A**] Independent causal estimates of T2D and genetically proxied raised HbA1c level on IPF risk. **[B]** Independent causal estimates of T2D and genetically proxied raised fasting insulin (F-insulin) level on IPF risk. **[C]** Independent causal estimates of T2D and genetically proxied raised BMI on IPF risk.

No causal effects were found of genetically proxied raised fasting insulin level on IPF risk (IVW-RE, OR = 0.60, 95% CI: 0.31-1.15, p = 0.124). Results of main MVMR analyses did not suggest any independent effects of T2D (MVMR-IVW, OR = 1.02, 95% CI: 0.94-1.12, p = 0.611) or raised fasting insulin (MVMR-IVW, OR = 0.69, 95% CI: 0.36-1.36, p = 0.285) on IPF risk. An independent protective effect of raised fasting insulin was indicated by MVMR-Egger results (OR = 0.37, 95% CI: 0.14-0.97, p = 0.044), however these results were not supported by MVMR-IVW, MVMR-Median, or MVMR-Lasso analyses so are unlikely to represent a genuine causal relationship (Figure 5B).

Estimates obtained by IVW-RE suggested a causal effect of raised BMI on IPF risk (IVW-RE, OR = 1.44, 95% CI: 1.12-1.85, p = 0.004), and MR-PRESSO did not suggest presence of pleiotropy (no outliers). The IVW-RE results were supported by MR-Lasso (OR = 1.35, 95% CI: 1.07-1.71, p = 0.011). MR estimates from weighted median (OR = 1.26, 95% CI: 0.87-1.81, p = 0.225) and MR-Egger methods (OR = 1.38, 95% CI: 0.71-2.70, p = 0.342) were concordant with IVW-RE but were not statistically significant, which may be due to the lower statistical power of these methods^44^. Results of MVMR analyses did not suggest any independent effects of T2D (MVMR-IVW, OR = 0.91, 95% CI: 0.68-1.21, p = 0.508) or raised BMI (MVMR-IVW, OR = 1.01, 95% CI: 0.94-1.09, p = 0.816) on IPF risk. Estimates obtained using pleiotropy-robust methods were consistent with those of MVMR-IVW (Figure 5C).

## Discussion

The results of this study suggest that DM is unlikely to be in the causal pathway of IPF, either independently (T1D, T2D) or through related factors (raised HbA1c and fasting insulin). A significant positive effect was suggested of BMI on IPF, but this effect did not persist in MVMR analyses of T2D on IPF with BMI as a secondary exposure.

Using T2D as an exposure in MR has limitations including instrumentation of a binary exposure based on a threshold of an underlying continuous variable, and potential confounding from related risk factors. To address these limitations additional two-sample MR analyses were performed with continuous exposures aiming to represent main factors associated with T2D – blood glucose (HbA1c level), dysregulated insulin (fasting insulin level), and obesity (BMI). The use of MR to investigate the relationship between T2D and IPF is limited by the complexity of both traits. The use of underlying continuous exposures and pleiotropy-robust methods, aimed to address this complexity. Further, a multivariable MR approach (MVMR) was performed to distinguish the independent effects of these correlated exposures (T2D, HbA1c, fasting insulin, and BMI). The inclusion of MVMR analyses of DM-associated factors therefore investigates potential causal pathways between DM and IPF in a more targeted and comprehensive approach. This study did not suggest causal effects through any of the investigated pathways, but interacting mechanisms between DM and IPF may have non-linear effects, which would not be represented in the MR approaches applied here. Further, causal effects across diseases may only exist in a subset of individuals or when in combination with additional genetic traits or environmental factors.

The lack of causal effects in either direction between T2D and IPF risk are consistent with the findings of another recent study^51^. Compared to this previous study, our investigation provides a more comprehensive assessment of possible causal pathways by targeting specific pathways between DM and IPF. Use of HbA1c instruments has been demonstrated to be superior to diabetes instruments in strength and variance explained^52^, and enables further insights into potential interacting disease mechanisms with less potential for confounding. Inclusion of HbA1c and T2D as exposures therefore strengthens our study. The null effects of raised HbA1c on IPF disease risk do not align with previously proposed mechanisms of systemic hyperglycemia promoting lung fibrosis through disrupted DNA repair and increased senescence^21,53^. Our study also included an MR analysis of T1D to assess possible effects specific to this form of DM.

The comorbid relationship between DM and IPF may result from shared factors affecting both traits, rather than directly causative effects. For example, T2D and IPF share risk factors such as smoking and ageing^54,55^. Results of univariable MR analyses suggested a positive effect of BMI on IPF risk, however no independent effects of T2D or BMI on IPF were indicated by MVMR results. A previous study has suggested that IPF patients are generally normally nourished or obese^56^, but lower BMI is associated with poorer disease outcomes^57,58^. Given the complexity of factors contributing to BMI, the pathway by which BMI could influence IPF risk is unclear. It is therefore difficult to rule out potential confounding driven by shared upstream factors and uncertain if any effect of BMI could underlie the association between DM and IPF. Alternatively, factors such as shortened telomere length may independently drive both IPF and T2D. Shorter telomere length, which has been shown to be a causative factor in IPF^59^, is associated with diabetes progression^60^, insulin resistance^61^ and increased HbA1c level^62,63^, obesity^64,65^, and telomere attrition is predicted to drive pancreatic beta cell dysfunction^66^. As telomere length also decreases with increased age and cigarette smoking^67,68^, this may also explain shared associations of these risk factors with T2D and IPF. Further, potential interaction of treatments across diseases may exist. IPF patients experiencing acute exacerbations are routinely treated with corticosteroids^69^, which promote hyperglycemia and diabetes^70,71^. Similarly, metformin, which is the first line oral therapy for the treatment of type 2 diabetes^72^, has been suggested to resolve lung fibrosis in cellular and animal models^73–75^ which may suggest the presence of shared disease pathways.

The findings of this study suggest that the comorbid relationship between DM and IPF is not driven by causative effects of either disease. Intrinsic mechanisms upstream of both traits and environmental effects may underlie this comorbid relationship. Shared mechanisms independently driving both traits may highlight opportunities for intervention and warrant further investigation.

## Data availability

Summary results for the GWAS of IPF risk were requested through the collaborative genetic studies of idiopathic pulmonary fibrosis GitHub page: (https://github.com/genomicsITER/PFgenetics). Summary data for all other GWAS studies were retrieved from GWAS catalog (https://www.ebi.ac.uk/gwas/).

## Supplement

### Supplementary analysis 1: An MR analysis of T2D on IPF using GWAS of European ancestry only

#### Methods

Main MR analyses used the largest disease-targeted GWAS of T2D by Xue *et al* (2018)^33^. A limitation of this dataset was the mixed ancestry cohort used, where the IPF GWAS cohort was European ancestry only. As sensitivity analysis, a two-sample MR analysis of T2D on IPF was performed using a European ancestry GWAS of T2D^38^ (12,931 European ancestry cases, 57,196 European ancestry controls).

#### Results

Results using European ancestry GWAS were consistent with main analyses, suggesting a null effect of T2D on IPF (IVW-RE, OR = 0.99, 95% CI: 0.89-1.11, p = 0.916) (Supplementary Figure 1)

### Supplementary analysis 2: An MR analysis of HbA1c on IPF risk with only functionally glycemic HbA1c genetic variants

#### Methods

A two-sample MR analysis was performed to assess for evidence of causal effects of HbA1c on IPF using only glycemic HbA1c variants (associated with glycemic traits, p < 0.0001). 12 independent genetic variants significantly (p<5×10^-8^) associated with HbA1c level were identified from a recent GWAS^35^ (sample size: 88,355 of European ancestry).

#### Results

Results of MR analyses of HbA1c (glycemic variants only) on IPF did not suggest that raised HbA1c causes IPF (IVW-RE, OR = 2.78, 95% CI: 0.97-8.02, p = 0.058). Results from pleiotropy-robust methods were consistent with IVW-RE estimates (Supplementary Figure 2).

### Supplementary analysis 3: Secondary analysis assessing for causal effects of IPF on T2D

#### Methods

A two-sample MR analysis was performed to assess for evidence of causal effects of IPF on T2D. Nineteen independent genetic variants significantly (p<5×10^-8^) associated with IPF risk were identified from a recent GWAS^34^ (4,125 IPF cases, 20,464 controls). Nine genetic instruments were found to be missing from the T2D GWAS study used in main analyses, with no available proxies (R^2^ > 0.7) shared across both exposure and outcome GWAS. Sensitivity MR analyses were performed using a different T2D GWAS study containing all 19 IPF variants^76^ (12,931 European ancestry cases, 57,196 European ancestry controls) to that.

#### Results

Results of MR analyses (9 IPF instruments) of IPF on T2D did not suggest that IPF causes T2D (IVW-RE, OR = 1.00, 95% CI: 0.97-1.03, p = 0.795). Results from pleiotropy-robust methods were consistent with IVW-RE estimates (Supplementary Figure 2). Results from sensitivity analyses using all 19 IPF variants as exposure instruments were consistent previous estimates using 9 IPF variants (IVW-RE, OR = 0.97, 95% CI: 0.93-1.01, p = 0.176) (Supplementary Figure 3)

### Supplementary analysis 4: Secondary analysis assessing for causal effects of IPF on T1D

#### Methods

A two-sample MR analysis was performed to assess for evidence of causal effects of IPF on T1D. 19 independent genetic variants significantly (p<5×10^-8^) associated with IPF risk were identified from a recent GWAS^34^ (4,125 IPF cases, 20,464 controls). 14 genetic instruments were found to be missing from the T2D GWAS study used in main analyses, with no available proxies (R^2^ > 0.7) shared across both exposure and outcome GWAS.

#### Results

Results of MR analyses (5 IPF instruments) of IPF on T1D did not suggest that IPF causes T1D (IVW-RE, OR = 1.11, 95% CI: 0.95-1.28, p = 0.184). Results from pleiotropy-robust methods were consistent with IVW-RE estimates (Supplementary Figure 4).

**Supplementary Figure 1:**
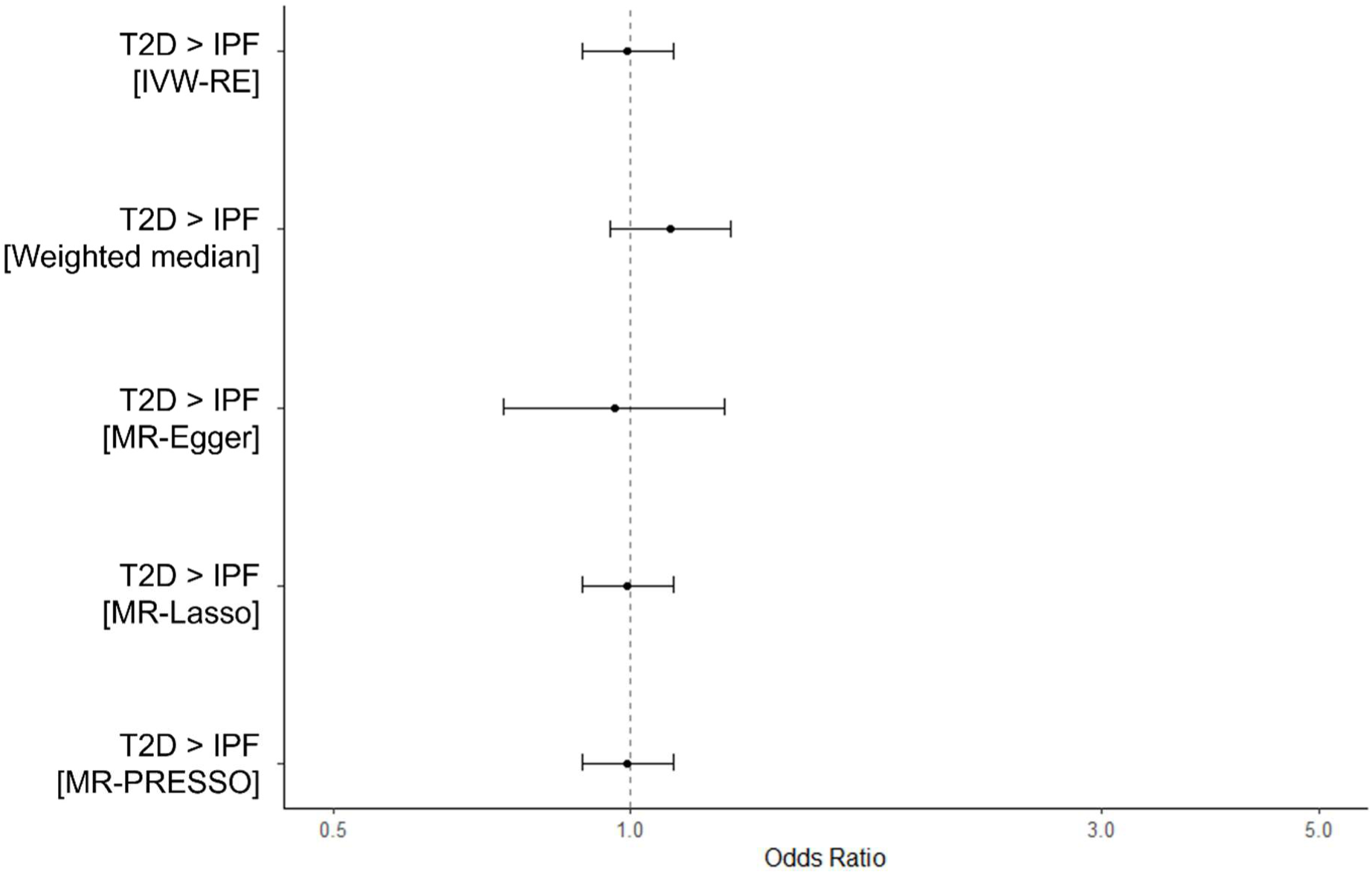
Overall causal estimates of T2D on IPF risk (Exposure GWAS cohort all of European ancestry)

**Supplementary Figure 2:**
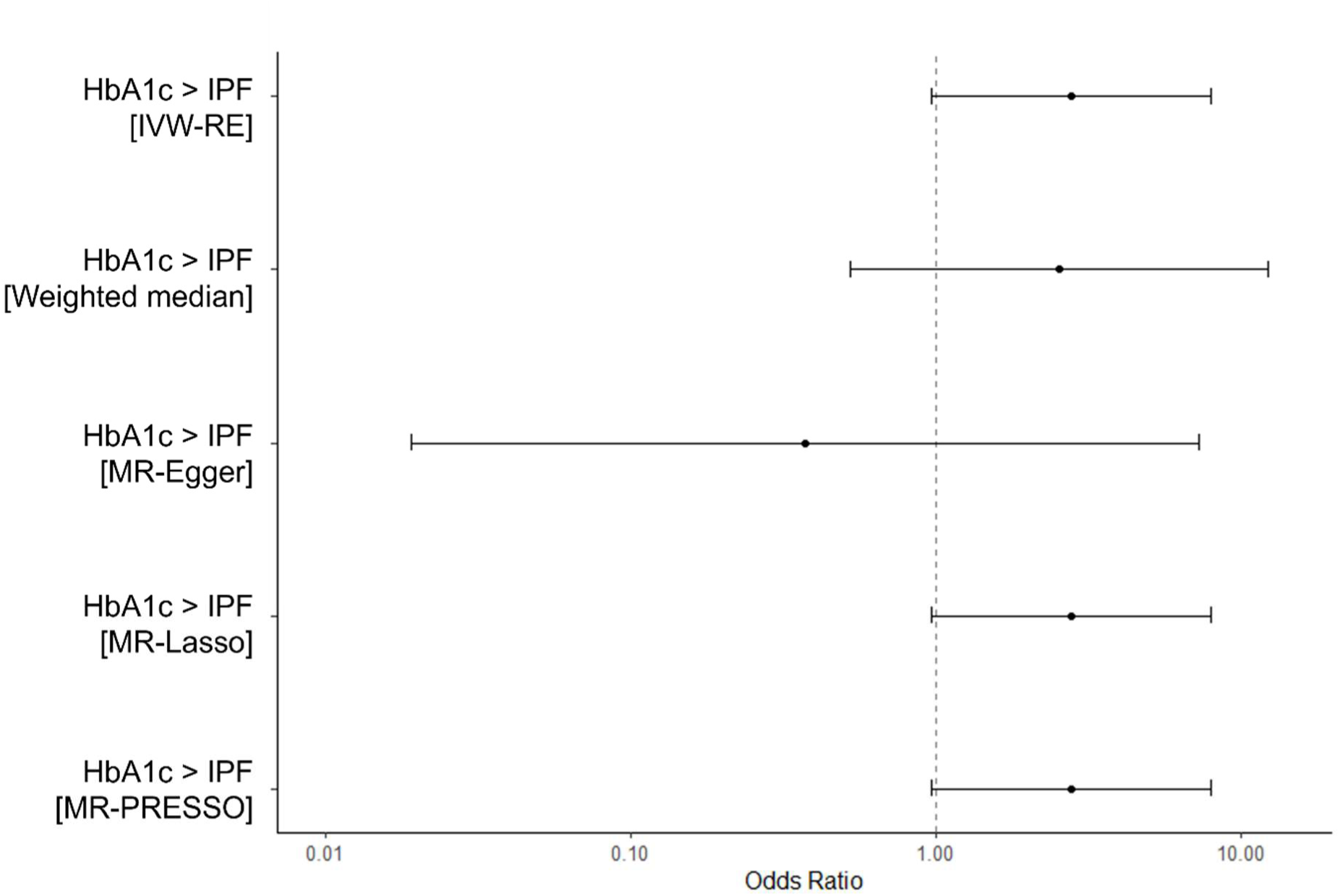
Overall causal estimates of HbA1c on IPF risk (only glycemic HbA1c variants)

**Supplementary Figure 3:**
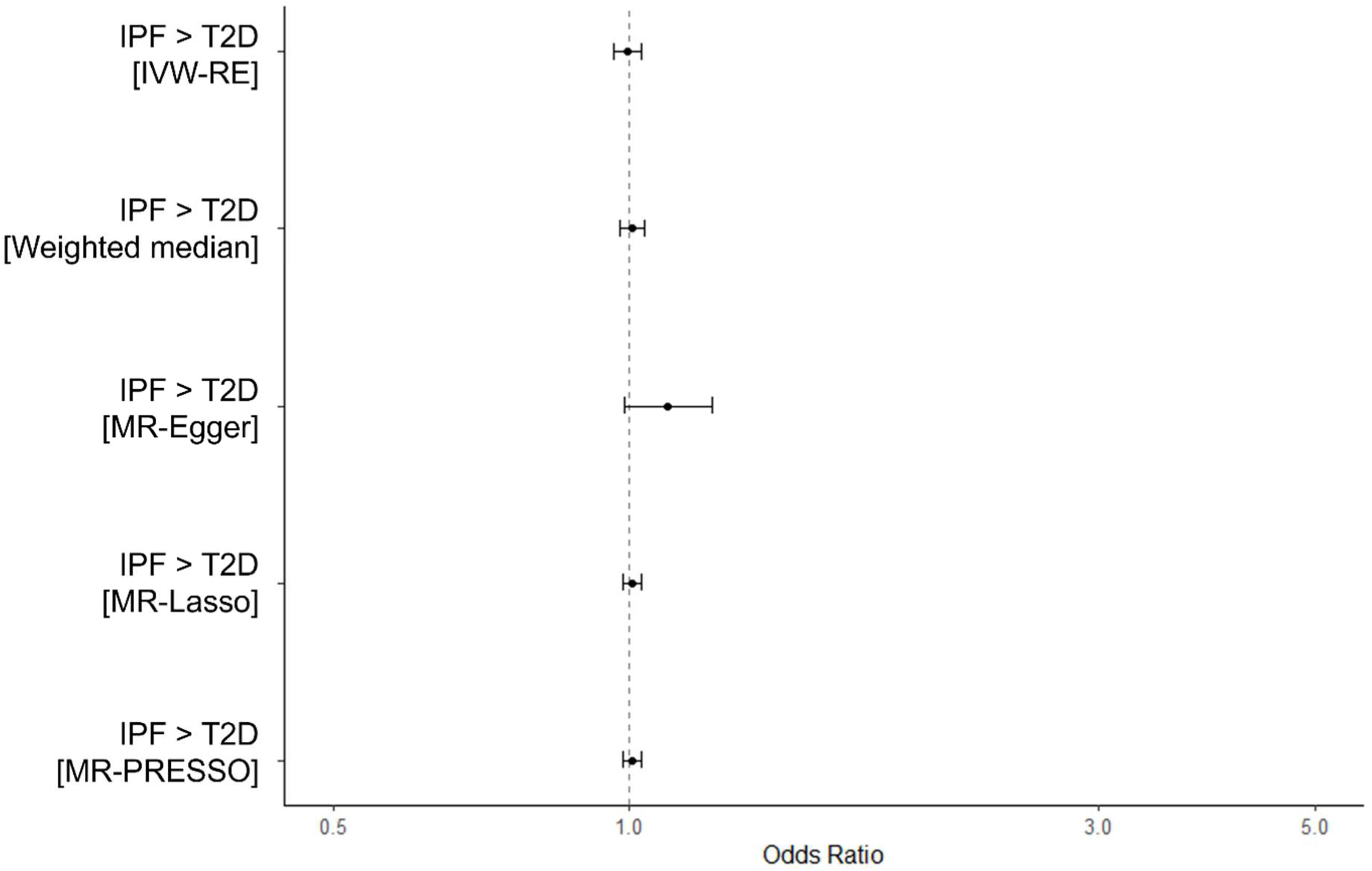
Overall causal estimates of IPF on T2D. 10/19 IPF variants were not present in the exposure (T2D) dataset for this analysis.

**Supplementary Figure 4:**
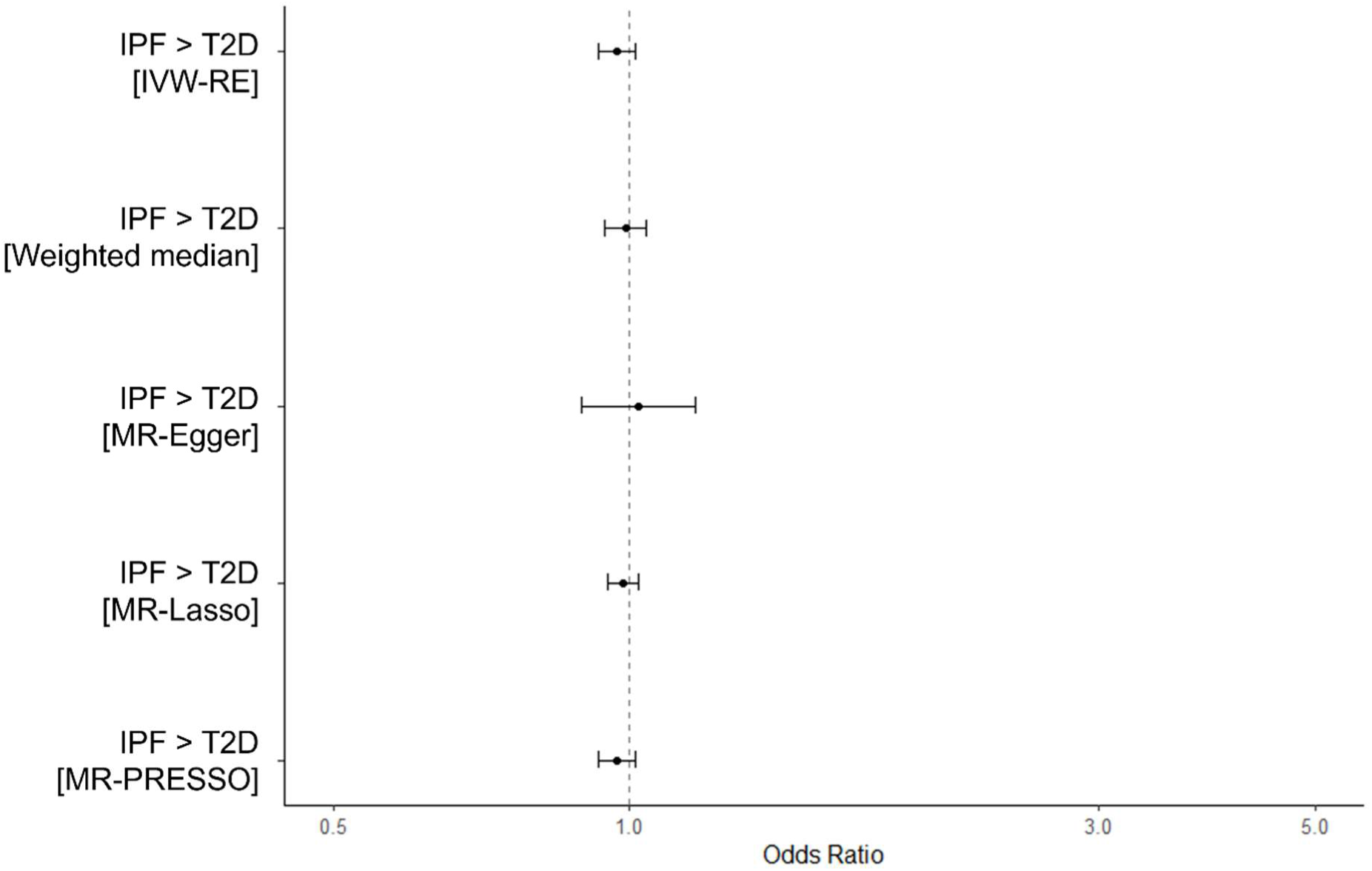
Overall causal estimates of IPF on T2D. Sensitivity analysis using a T2D GWAS with only European ancestry and all 19 IPF variants present in the outcome dataset.

**Supplementary Figure 5:**
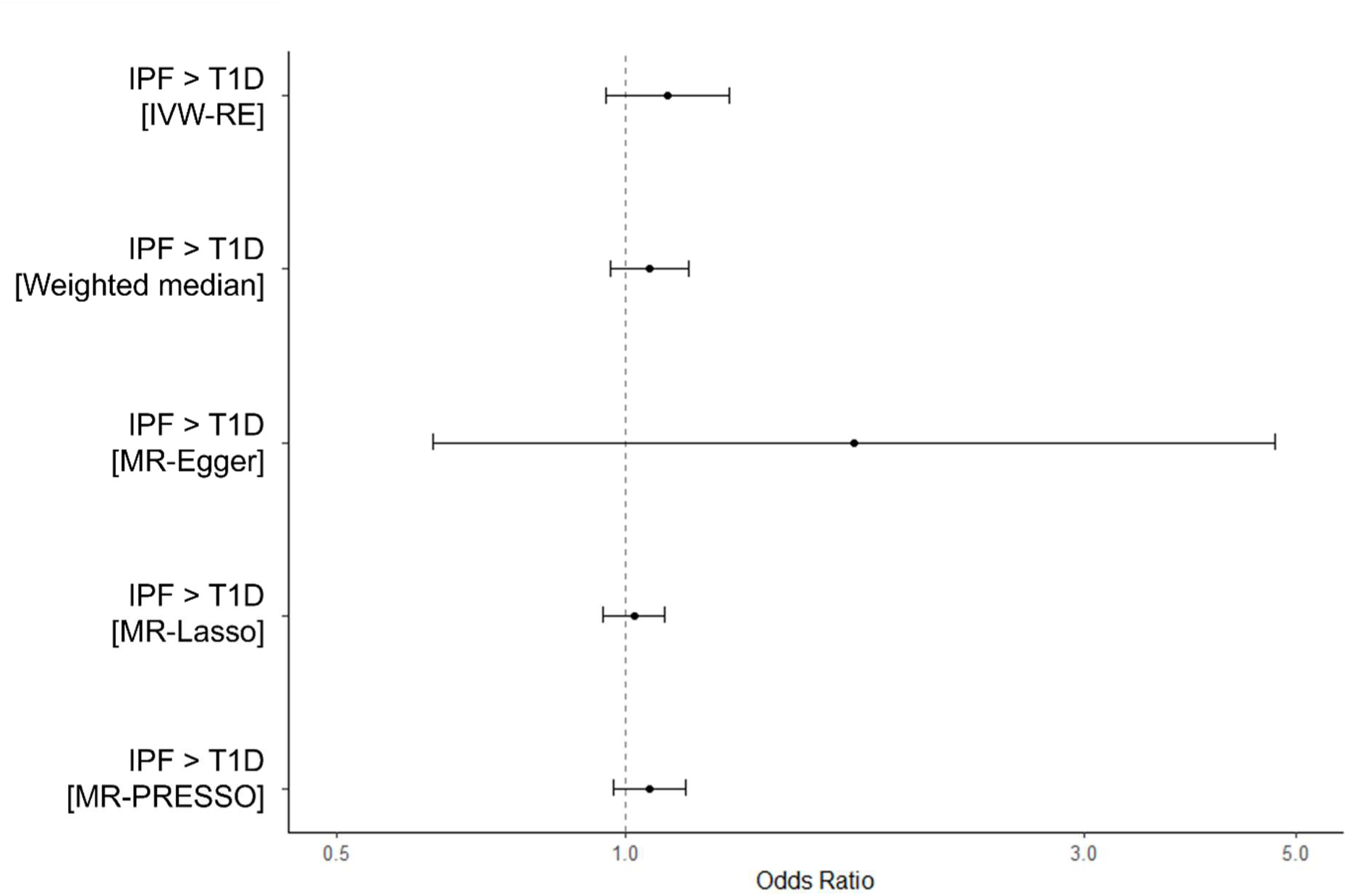
Overall causal estimates of IPF on T1D.

**Supplementary Figure 6:**
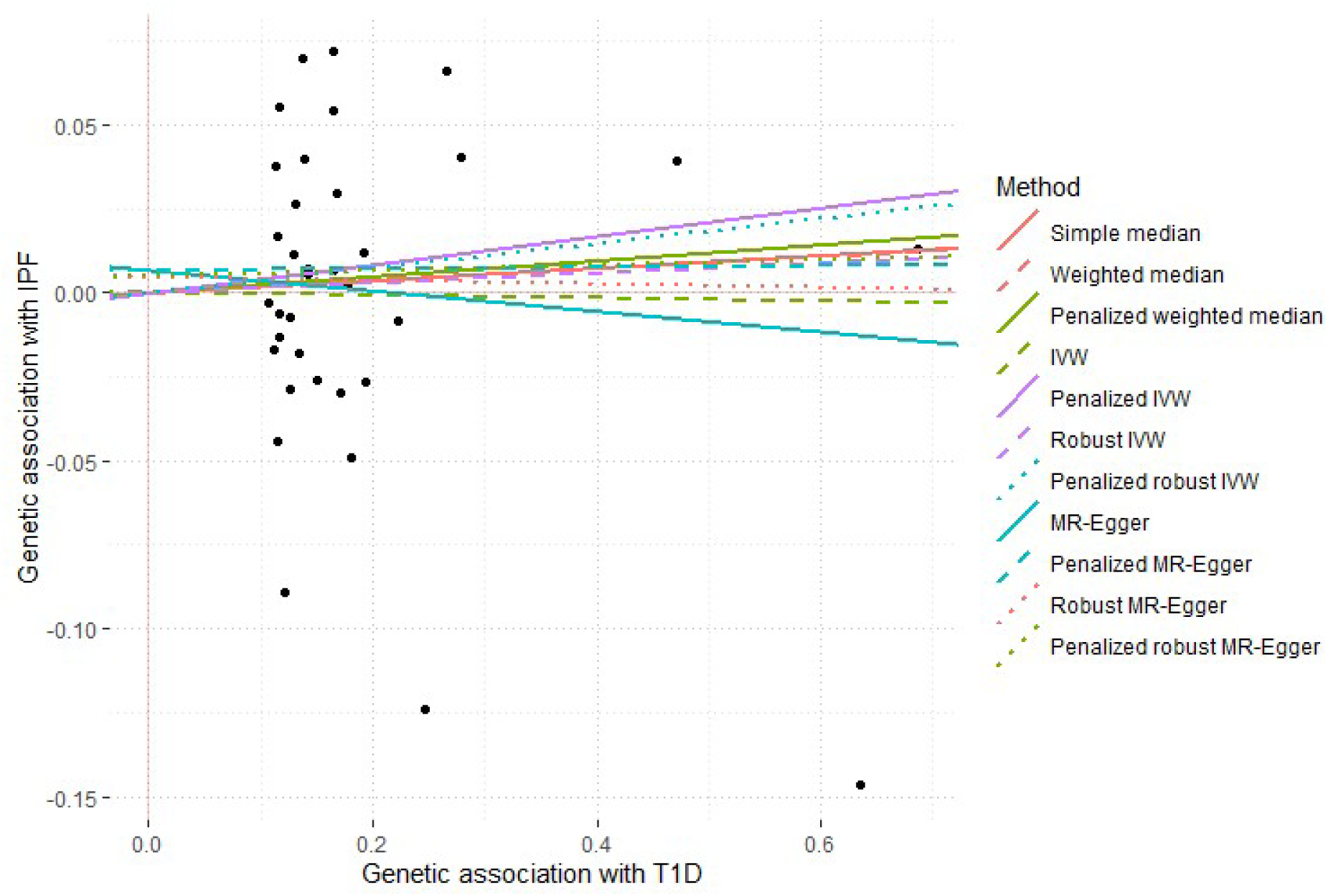
A scatter plot of single variant effect estimates on T1D (exposure) (x-axis) and IPF (outcome) (y-axis) and overall MR meta-analysis estimates from different methods. Genetic association statistics are beta effect coefficients derived from GWAS summary statistics for each trait.

**Supplementary Figure 7:**
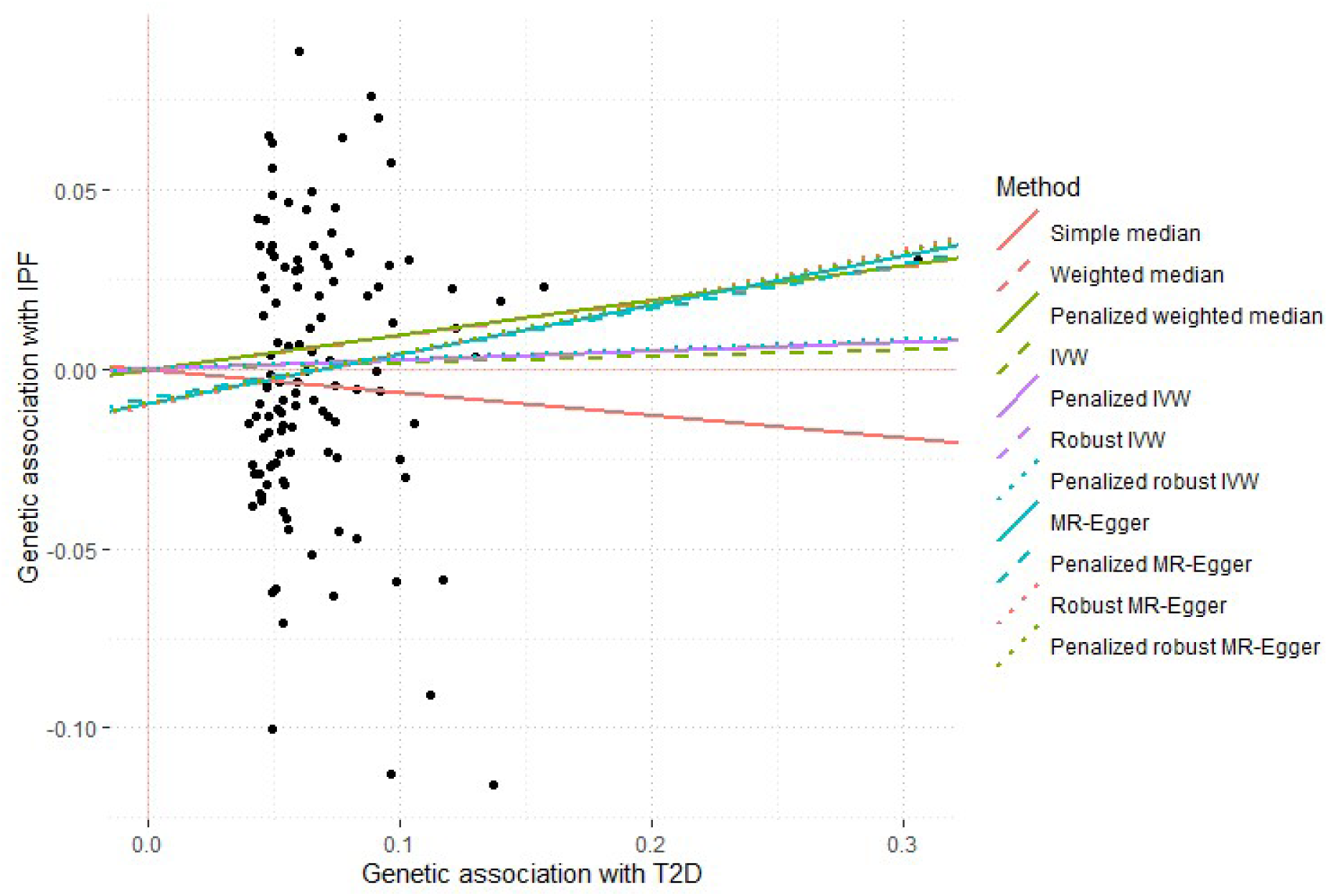
A scatter plot of single variant effect estimates on T2D (exposure) (x-axis) and IPF (outcome) (y-axis) and overall MR meta-analysis estimates from different methods. Genetic association statistics are beta effect coefficients derived from GWAS summary statistics for each trait.

**Supplementary Figure 8:**
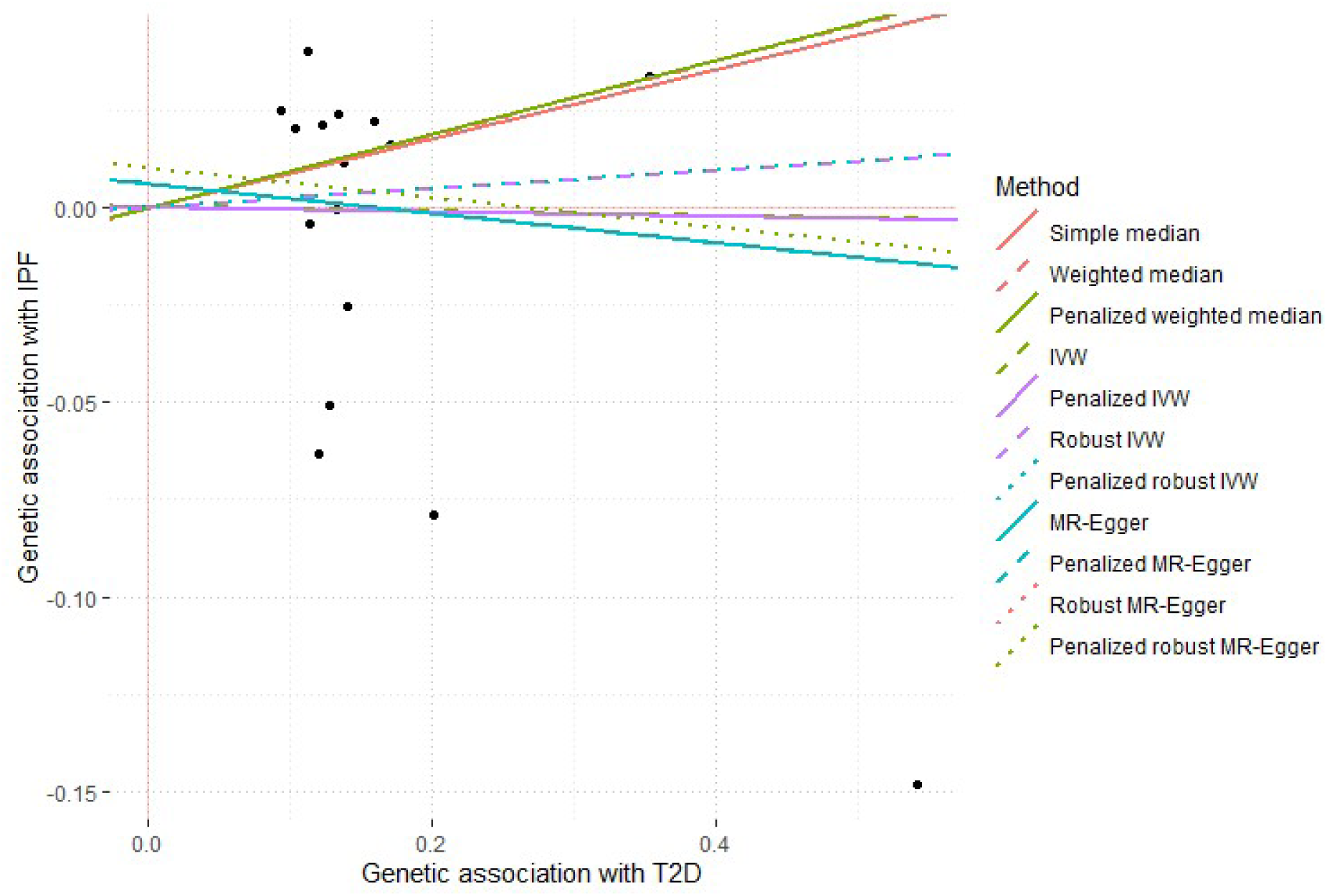
A scatter plot of single variant effect estimates on T2D (European ancestry only) (exposure) (x-axis) and IPF (outcome) (y-axis) and overall MR meta-analysis estimates from different methods. Genetic association statistics are beta effect coefficients derived from GWAS summary statistics for each trait.

**Supplementary Figure 9:**
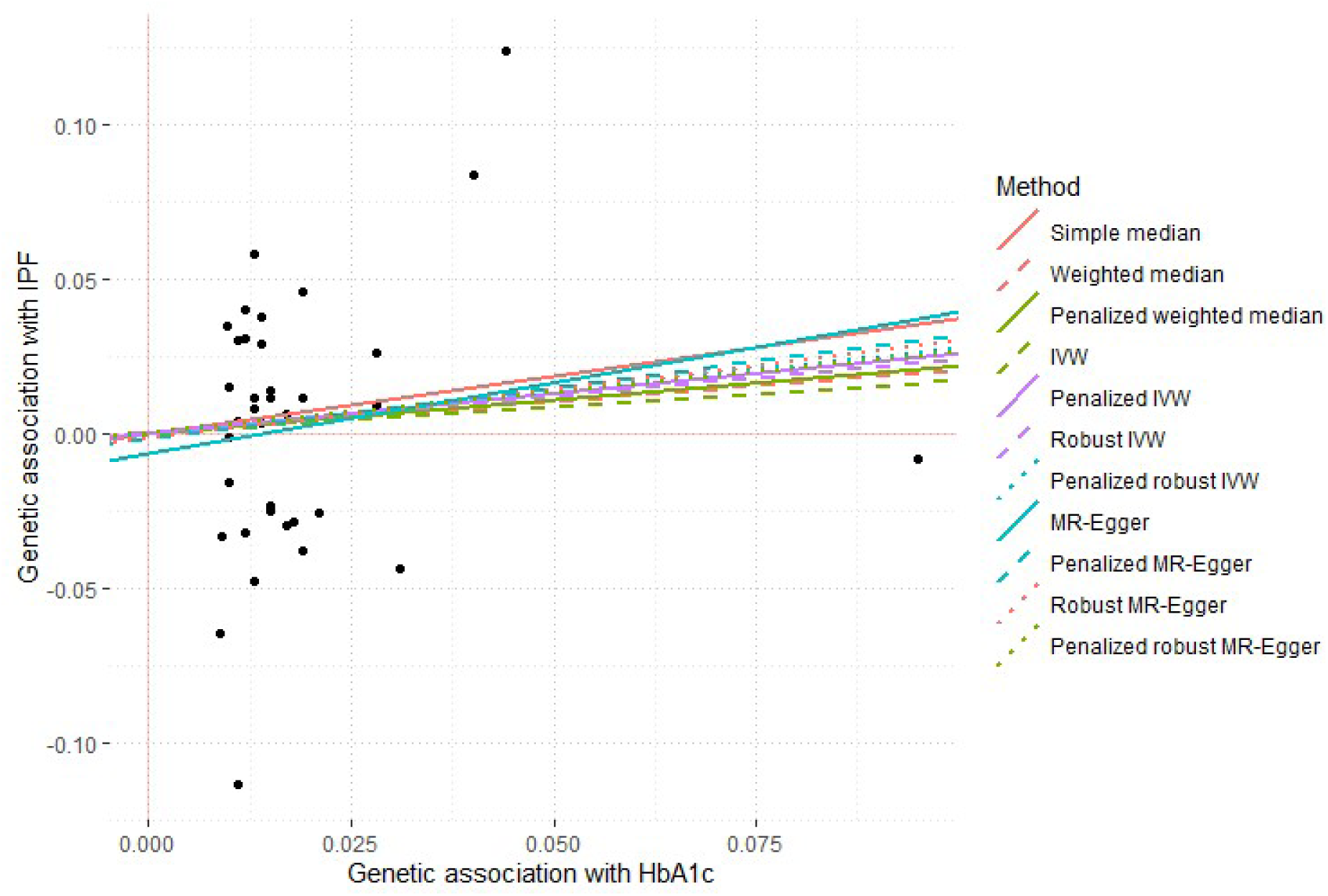
A scatter plot of single variant effect estimates on HbA1c (exposure) (x-axis) and IPF (outcome) (y-axis) and overall MR meta-analysis estimates from different methods. Genetic association statistics are beta effect coefficients derived from GWAS summary statistics for each trait.

**Supplementary Figure 10:**
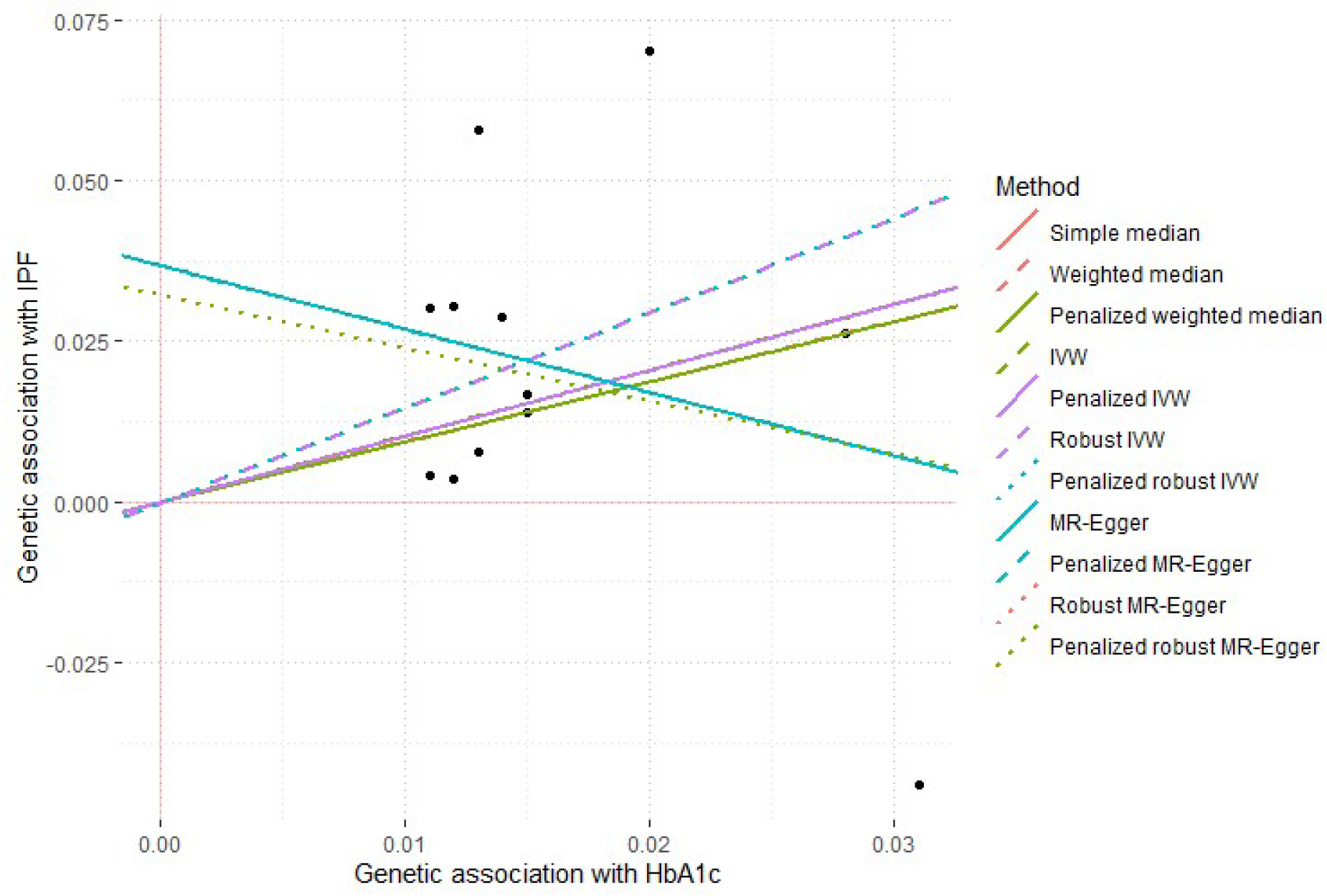
A scatter plot of single variant effect estimates on HbA1c (glycemic genetic variants only) (exposure) (x-axis) and IPF (outcome) (y-axis) and overall meta-analysis estimates from different methods. Genetic association statistics are beta effect coefficients derived from GWAS summary statistics for each trait.

**Supplementary Figure 11:**
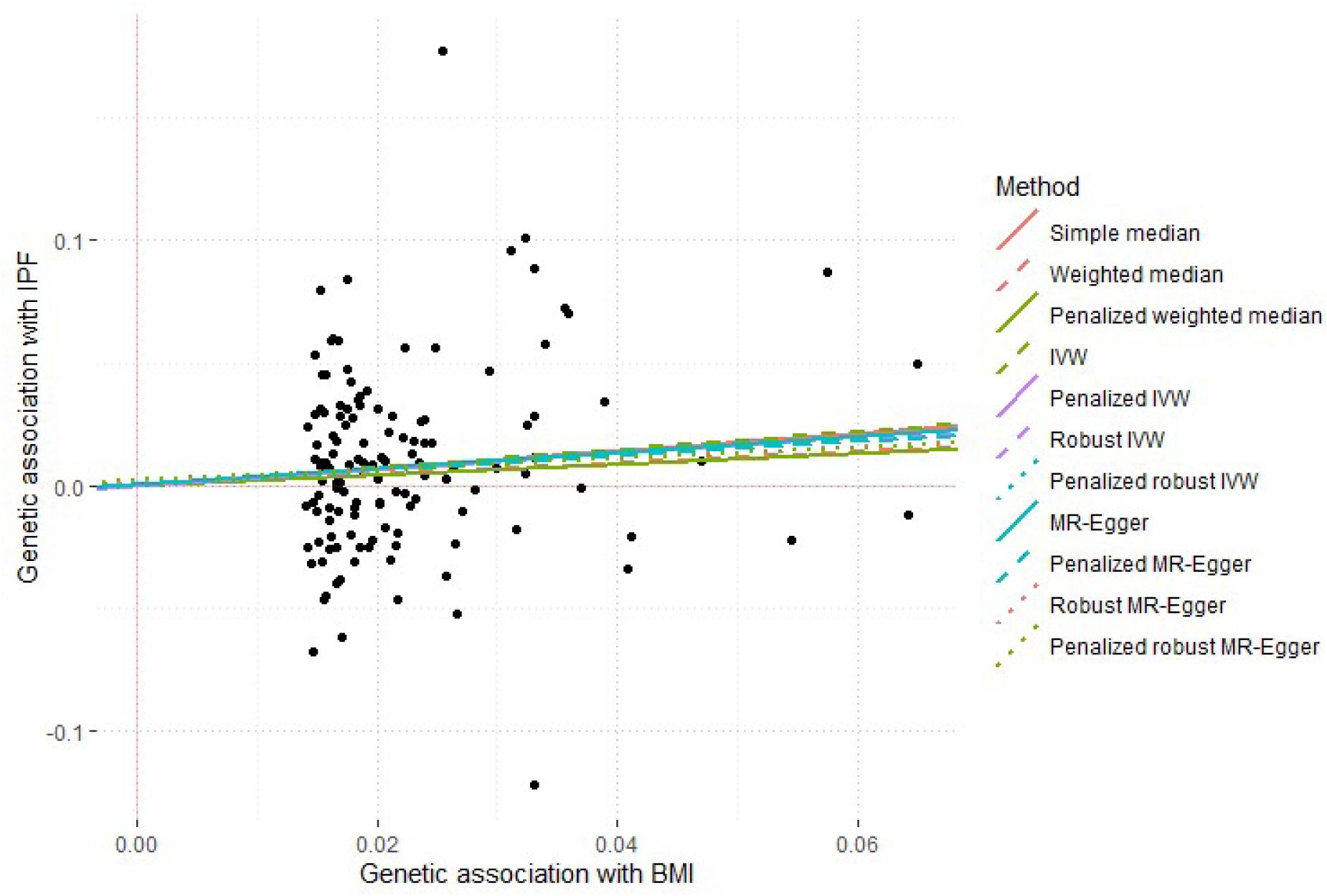
A scatter plot of single variant effect estimates on BMI (exposure) (x-axis) and IPF (outcome) (y-axis) and overall meta-analysis estimates from different methods. Genetic association statistics are beta effect coefficients derived from GWAS summary statistics for each trait.

**Supplementary Figure 12:**
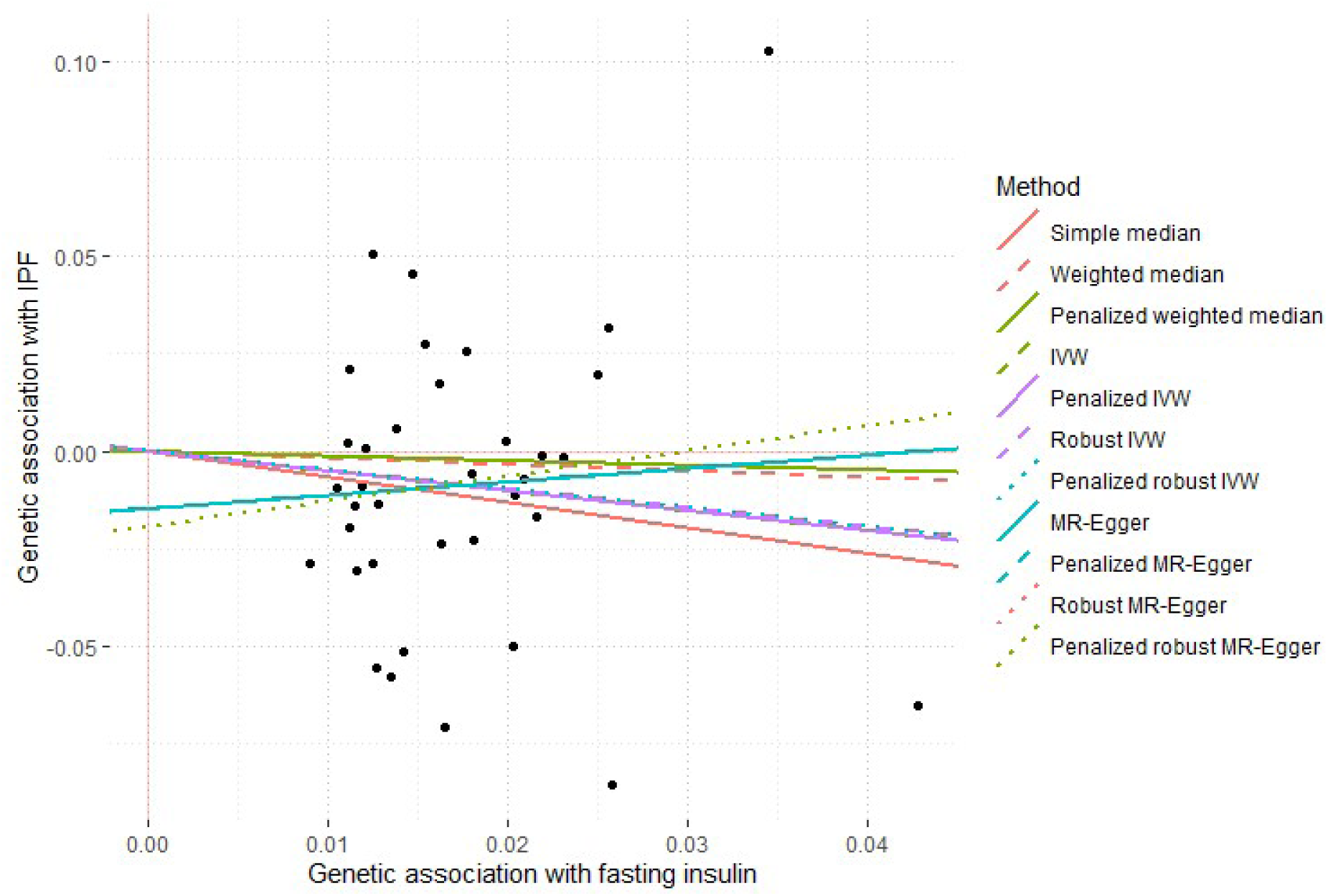
A scatter plot of single variant effect estimates on fasting insulin (exposure) (x-axis) and IPF (outcome) (y-axis) and overall meta-analysis estimates from different methods. Genetic association statistics are beta effect coefficients derived from GWAS summary statistics for each trait.

**Supplementary Figure 13:**
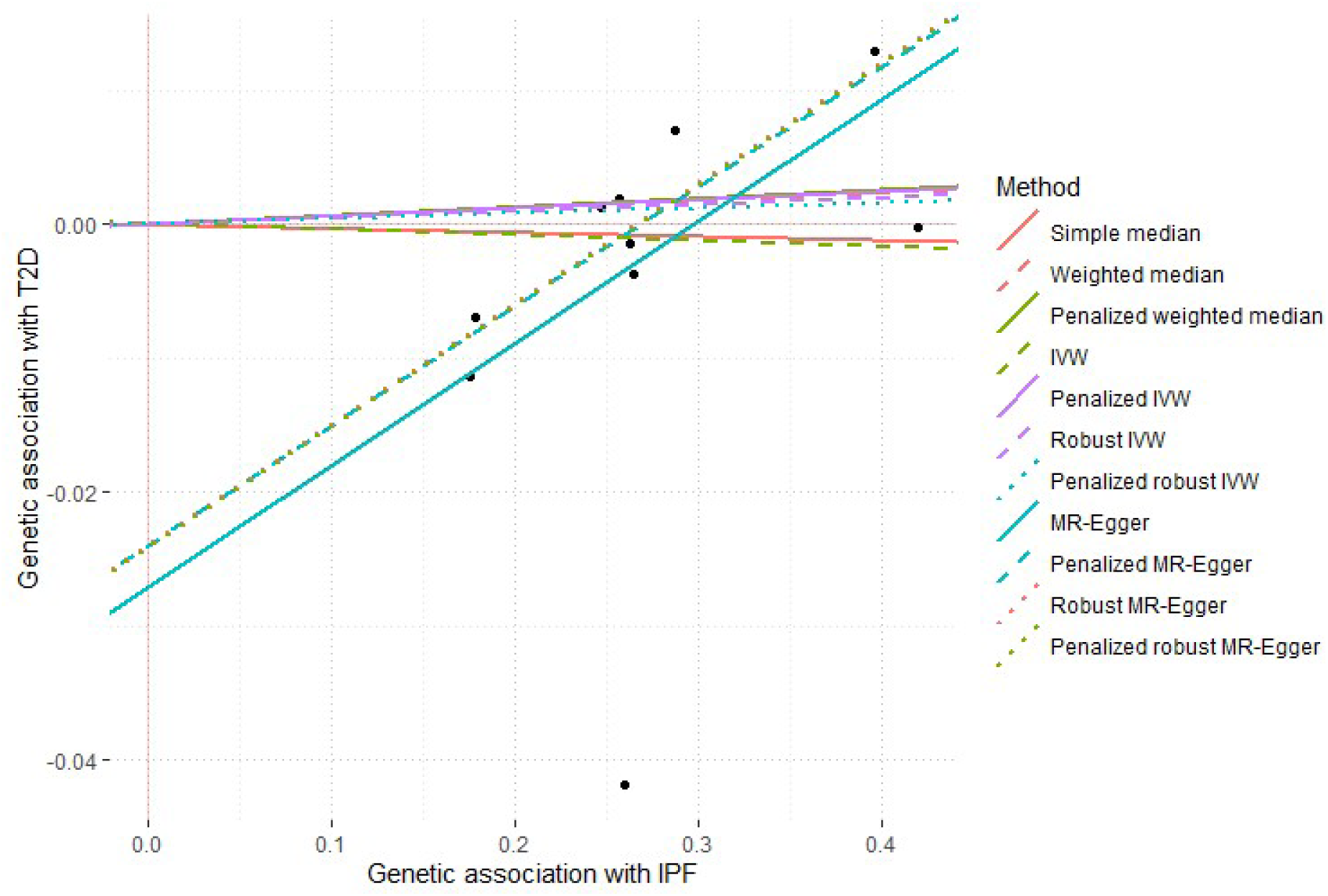
A scatter plot of single variant effect estimates on IPF (exposure) (x-axis) and T2D (outcome) (y-axis) and overall meta-analysis estimates from different methods. Genetic association statistics are beta effect coefficients derived from GWAS summary statistics for each trait.

**Supplementary Figure 14:**
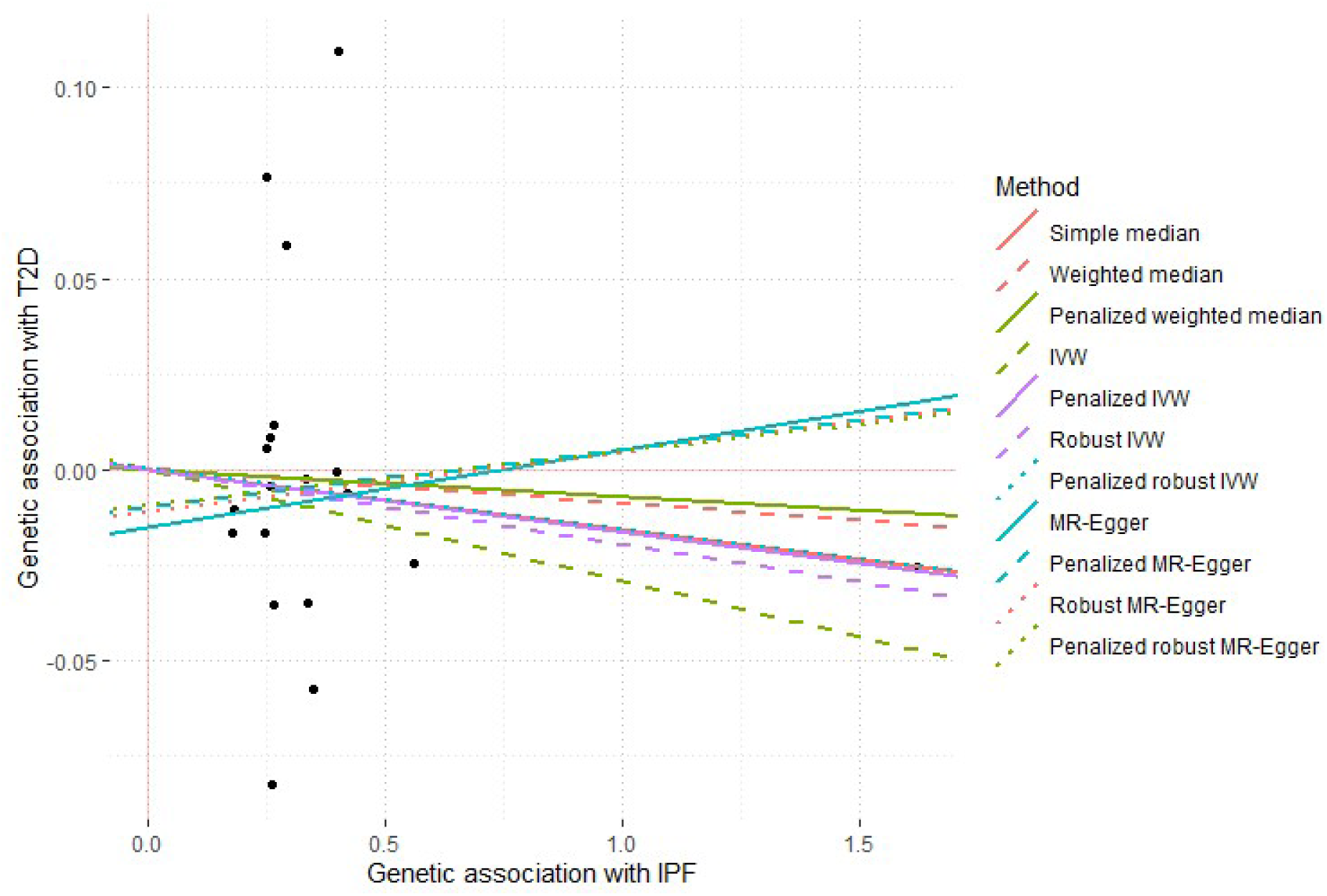
A scatter plot of single variant effect estimates on IPF (exposure) (x-axis) and T2D (outcome) (y-axis) (European ancestry only, containing all 19 IPF variants) and overall meta-analysis estimates from different methods. Genetic association statistics are beta effect coefficients derived from GWAS summary statistics for each trait.

**Supplementary Figure 14:**
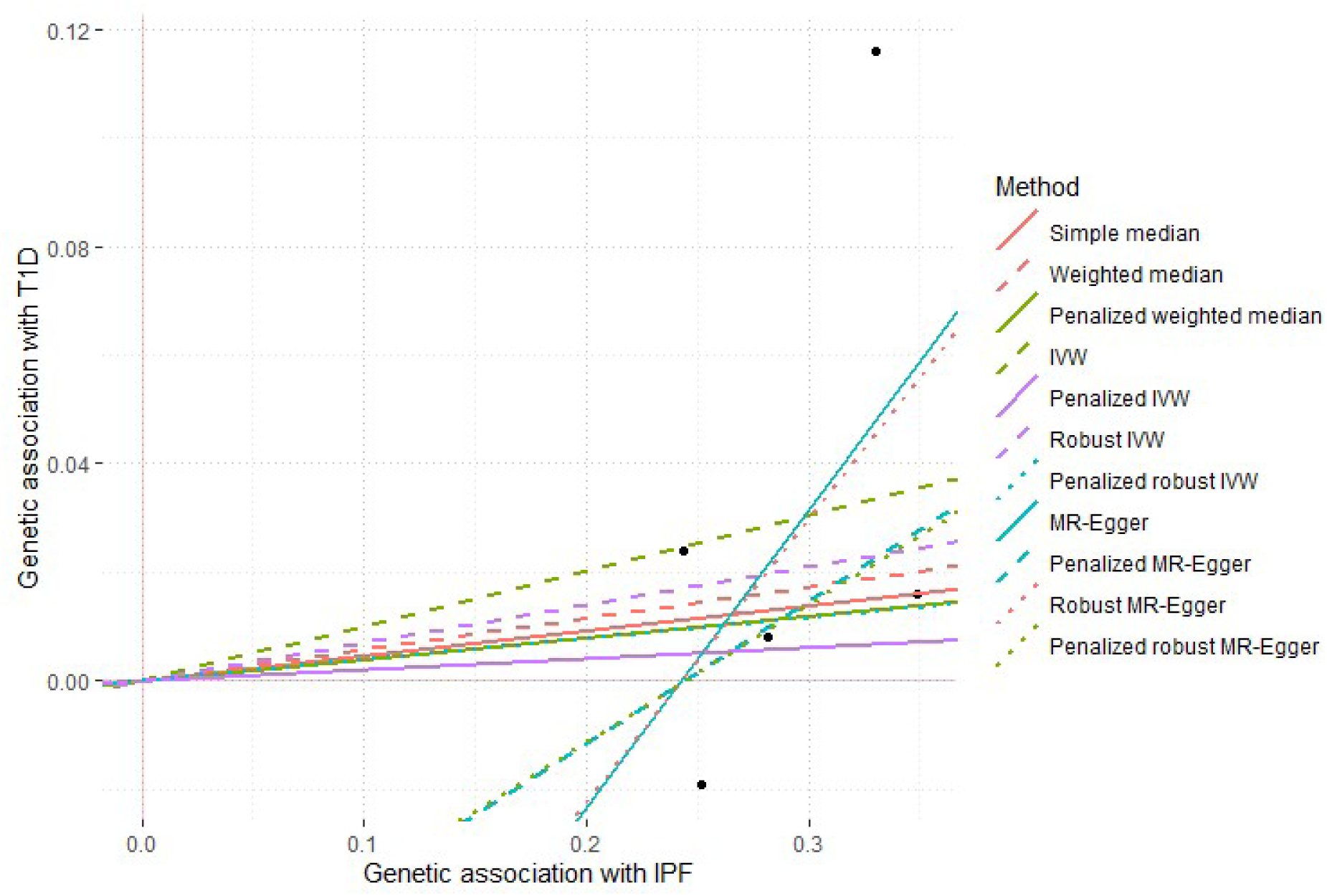
A scatter plot of single variant effect estimates on IPF (exposure) (x-axis) and T1D (outcome) (y-axis) and overall meta-analysis estimates from different methods. Genetic association statistics are beta effect coefficients derived from GWAS summary statistics for each trait.

